# Retained Surgical Item Incidence in the United States from 2016 to 2023: A Descriptive Study

**DOI:** 10.1101/2025.06.26.25329866

**Authors:** Danel Mayan, Pam Kumparatana, Agith Antony, Kenneth A. Taylor

## Abstract

**Importance:** Retained surgical items (RSI) are serious, preventable medical errors that continue to occur despite widespread implementation of safety protocols and technological interventions. Understanding RSI incidence can inform quality improvement aimed at prevention and patient safety.

**Objective:** To estimate the incidence of retained surgical items in the United States from 2016 to 2023 overall, annually, and stratified by state and surgery category.

**Design:** This was a longitudinal retrospective cohort study utilizing de-identified healthcare claims data from 2016 to 2023.

**Setting:** Data were from the Komodo Research Dataset, which aggregates medical and pharmacy claims from US commercial and government insurance sources.

**Participants:** 198,619,904 surgeries between January 1, 2016, and December 31, 2023 where the patient had at least six months of prior continuous medical and pharmacy insurance enrollment. Surgeries were excluded if there was evidence of RSI before surgery or present on admission, surgery was performed in a US territory, or the patient was aged < 2 on the surgery date.

**Exposure:** Surgical procedures identified by administrative codes categorized as broad or narrow by the Agency for Healthcare Research and Quality.

**Main Outcome(s) and Measure(s):** The primary outcome was the incidence proportion of RSIs, identified using International Classification of Diseases, Tenth Revision, Clinical Modification codes up to 365 days after surgery. Incidence was estimated overall, annually, and stratified by state and surgical category. Data were analyzed from March 2025 to May 2025.

**Results:** The overall RSI incidence was 1.34 (95% CI: 1.32 to 1.36) per 10,000 surgeries, decreasing from 1.63 (95% CI: 1.58 to 1.70) in 2016 to 1.08 (95% CI: 1.04 to 1.12) in 2023. The 5 highest state-stratified RSI incidences were in the District of Columbia, Maine, Nebraska, Ohio, and West Virginia. Exploratory laparotomies and transplant surgeries had the highest surgery category-stratified RSI incidence.

**Conclusion and Relevance:** This large national claims-based study estimated RSI incidence and describes associated patterns in the US, confirming a persistent but declining annual incidence across the 8-year period. These findings highlight the ongoing need for targeted prevention strategies and system-wide efforts to reduce RSIs and improve surgical care.

**Key Points:** *Question:* What is the incidence of retained surgical items from 2016 to 2023 in the United States overall, annually, and stratified by surgery type and geographic region?

*Findings:* In this descriptive study of over 198 million surgical procedures from 2016 to 2023, retained surgical item incidence was 1.34 per 10,000 surgeries, declining from 1.63 in 2016 to 1.08 per 10,000 surgeries in 2023. The highest surgery category-stratified incidence was exploratory laparotomy; geographically-stratified incidence was highest in the District of Columbia.

*Meaning:* Retained surgical items remain a national issue, though declining, highlighting the need for targeted clinical and hospital prevention.

## Introduction

Retained surgical items (RSIs), defined as objects unintentionally left inside a patient after a surgical procedure, represent a significant, preventable threat to patient safety in the surgical setting.^1^ Despite advancements in surgical techniques and implementation of stringent safety protocols, RSIs continue to occur, leading to increased morbidity, extended hospital stays, additional surgical interventions, and heightened healthcare costs.^2–4^ Notably, RSIs have consistently ranked among the top sentinel events reported to The Joint Commission since the organization began publishing data in 2012.^5^

RSI incidence is challenging to estimate due to underreporting and variability in detection methods; however, existing literature provides some estimates.^6–8^ A systematic review encompassing 21 studies estimated RSI incidence at 1.3 per 10,000 surgical procedures, not specific to a particular surgery type.^9^ Another study focusing on major abdominal and pelvic surgeries estimated similar incidence between 2007 and 2011.^2^ Notably, certain high-risk procedures, such as emergency abdominal operations, have RSI incidence as high as 1 in 700 surgical cases.^10^

The consequences of RSIs are profound, encompassing severe complications such as infections, organ perforation, obstruction, and chronic pain.^11,12^ These adverse events typically necessitate additional surgical interventions while negatively impacting patient quality of life. Case-specific factors contributing to RSIs include procedural complexity, emergency case status, and patient comorbidities such as elevated body mass index.^4^ Environmental factors within the operating room have also been identified as contributors, such as failure to perform accurate sponge and instrument counts and other deviations from standard protocols.^4^

Accurate assessment of RSI incidence is limited by traditional detection methods like manual instrument counts, which are prone to error, and variability in institutional documentation practices and reporting mechanisms.^13^ Additionally, many existing studies on RSIs rely on data from individual hospital systems or electronic health records, which can be limited by underreporting and smaller sample sizes restricted to specific institutions or regions.^6–8^ Healthcare claims data offer a broader, more comprehensive view by capturing diverse, real-world encounters across populations and regions, enabling better estimation of RSI incidence and trends.

In this study, we aimed to leverage large-scale claims data to establish a more precise and comprehensive estimate of RSI incidence across a broad surgical population. In addition to quantifying incidence, we sought to identify patient-level characteristics and procedural patterns associated with increased RSI risk. The insights from this work have the potential to inform evidence-based policy recommendations, guide the development of targeted surgical safety interventions, and support broader efforts to reduce the incidence of RSIs.

## Methods

### Data Source and Study Design

We conducted a longitudinal retrospective cohort study to estimate RSI incidence using de-identified healthcare claims data from June 1, 2015 to December 31, 2024 (see **eFigure 1**) from the Komodo Research Dataset (Komodo Health, Inc.; San Francisco, CA), a real-world dataset that aggregates medical claims across commercial and government insurance sources in the United States. The Komodo Research Dataset is built on the Komodo Healthcare Map®, which includes longitudinal patient medical and pharmacy claims data with associated diagnostic, procedural, and prescription codes. Inpatient hospitalization claims data also include Medicare Severity Diagnosis Related Group (MS-DRG) codes. For inpatient hospitalization claims missing an MS-DRG code, Komodo Health® imputes the associated MS-DRG code using patient information, diagnosis codes, and procedures codes.^14^ The Komodo Health data used for this study is de-identified using the Expert Determination de-identification standard as specified by the Health Insurance Portability and Accountability Act of 1996 for Komodo’s Healthcare Map to protect against the risk of re-identification.^15^ Neither the investigators nor Komodo Health have a keycode or crosswalk that can be used to re-identify patients. The use of such de-identified data is not considered human subjects research and does not require ethics approval.

### Study Population and Exposure Definition

We estimated procedure-level incidence, identifying surgical events between January 1, 2016 and December 31, 2024 using Current Procedural Terminology codes categorized as narrow (major therapeutic) or broad (major diagnostic or minor therapeutic) surgical procedures as defined by the Agency for Healthcare Research and Quality (AHRQ)^16^ and a subset of relevant surgical discharge MS-DRG codes (also from AHRQ; see **eTable 1**).^17^ Surgical procedures were grouped into clinical categories using AHRQ’s clinical classification definitions for services and procedures and further categorized into more specific surgical categories (see **eTable 2**).^18^ We excluded the “other therapeutic procedures” category because the two procedures it included were too broad to be reliably assigned to a single clinical category. Procedures occurring on the same day for a given patient were counted once for every unique procedure code. We excluded surgery events with < 183 days of pre-surgery continuous medical and pharmacy enrollment (allowable gaps ≤ 45 days) to ensure a minimum amount of available pre-surgery data. We also excluded surgical events with evidence of previous RSI within 183 days to ensure surgery was not related to RSI removal. Surgeries flagged using surgical discharge MS-DRG codes were excluded if an RSI code was listed as present on admission. We further excluded procedures performed in US territories, limiting our study to states and the District of Columbia (DC). Finally, we excluded patients aged < 2 years at the time of surgery because claim dates for newborn infants are reduced to year only as part of Komodo Health’s data redactions for de-identification. This redaction would impact the accuracy of data ascertained during our 6-month look-back period for surgeries occurring at age < 2 years and preclude our ability to determine the ordering of events in newborn infants (age < 1 year) altogether.

### Patient and Hospital Characteristics

We used the 183-day pre-surgery period to capture comorbidities for the Elixhauser Comorbidity Index, which were used to derive a single index score.^19,20^ Patient demographic characteristics (age, gender, race/ethnicity, payer type, Census-based geographic region) and hospital characteristics (state and type) were defined on the date of surgery. We describe hospital characteristics ascertainment in the **eMethods**.

### Outcome Ascertainment

We assessed each eligible surgical procedure for detection of incident RSI for up to 365 days or end of continuous enrollment after each eligible surgical procedure using a predefined list of International Classification of Diseases, Tenth Revision, Clinical Modification codes.^21^ RSI codes occurring after surgery date are an indicator of delayed RSI detection; therefore we considered the RSI event date as the surgery date rather than the RSI detection date. If an RSI was detected within 365 days after multiple surgeries, it was assigned to the most recent surgical event date. After ascertaining incident RSIs, we restricted our analyses to surgical procedures that occurred between January 1, 2016 and December 31, 2023. Surgical events in 2024 were excluded to allow up to 365 days to detect incident RSIs, giving sufficient time for RSI detection resulting from both early (e.g., infection) and delayed (e.g., granulomas, adhesions) symptoms.^22,23^

### Statistical Analysis

We calculated incidence as the incidence proportion per 10,000 surgeries using all eligible surgical procedures from 2016 to 2023 (denominator) and related RSIs detected within the 365 day follow-up period (numerator). We also estimated annual incidence, as well as the state (based on hospital location) and surgery category-stratified incidence. If an individual had multiple surgeries occurring on the same date in different surgery categories, then all procedures for the individual on that date (and any associated RSI) were excluded from the surgery category-stratified incidence estimates to avoid misattributing RSI events to the wrong surgery category. We estimated 95% confidence intervals (CI) for all incidence proportion estimates using the Wilson score interval method.^24^ We calculated time to RSI detection as the difference (in days) between the surgery date and the RSI detection date. Data cleaning and manipulation was done using data build tool with structured query language; analyses were performed using R.^25–30^

## Results

After applying eligibility criteria, our dataset consisted of 198,619,904 surgical procedures (see **eFigure 2**) from 74,446,465 unique patients. **Table 1** shows demographic characteristics overall and by RSI outcome status. The distribution of demographic characteristics among RSI cases were proportional to the distribution of all eligible surgical cases with regard to age, sex, race/ethnicity, and geographic region of the procedure. Insurance type distribution did not show the same close proportionality. Commercially-insured patients represented 51.5% of procedures involved, but only 44.6% of RSI cases. Additionally, 24.3% of all procedures - but 30.3% of all RSI cases - occurred among Medicaid beneficiaries. RSI cases also appeared to disproportionately occur at academic hospitals, representing 41.1% of all RSI cases but only 28.2% of eligible procedures. In contrast, 61.4% of all surgeries occurred at community hospitals, but 52.0% only of all RSI cases. Demographic characteristics stratified by surgery category are presented in **eTable 3**.

**Table 1.** Pooled Demographic and Hospital Characteristics Overall and by Retained Surgical Item Status, 2016 to 2023.

| Characteristic | Overall,<br>N = 198,619,904 | No RSI,<br>N = 198,593,299 | RSI,<br>N = 26,605 |
| --- | --- | --- | --- |
| Age, y |  |  |  |
| Mean (SD) | 49.3 (20.0) | 49.3 (20.0) | 49.9 (18.8) |
| Median (IQR) | 53 (34, 64) | 53 (34, 64) | 52 (36, 63) |
| Min, Max | 2, 87 | 2, 87 | 2, 87 |
| Unknown | 396,735 | 396,688 | 47 |
| Age Group, No. (%) |  |  |  |
| < 18 | 14,846,610 (7.47) | 14,845,245 (7.48) | 1,365 (5.13) |
| 18-30 | 24,946,427 (12.6) | 24,943,120 (12.6) | 3,307 (12.4) |
| 31-40 | 24,898,103 (12.5) | 24,894,418 (12.5) | 3,685 (13.9) |
| 41-50 | 26,883,665 (13.5) | 26,879,679 (13.5) | 3,986 (15.0) |
| 51-60 | 40,873,004 (20.6) | 40,867,378 (20.6) | 5,626 (21.1) |
| 61-70 | 36,679,366 (18.5) | 36,674,468 (18.5) | 4,898 (18.4) |
| 71-80 | 20,624,189 (10.4) | 20,621,478 (10.4) | 2,711 (10.2) |
| ≥ 81 | 8,471,805 (4.27) | 8,470,825 (4.27) | 980 (3.68) |
| Unknown | 396,735 (0.20) | 396,688 (0.20) | 47 (0.18) |
| Sex, No. (%) |  |  |  |
| Female | 115,491,094 (58.1) | 115,475,920 (58.1) | 15,174 (57.0) |
| Male | 80,400,633 (40.5) | 80,389,581 (40.5) | 11,052 (41.5) |
| Unknown | 2,728,177 (1.37) | 2,727,798 (1.37) | 379 (1.42) |
| Insurance Type, No. (%) |  |  |  |
| Commercial | 102,302,362 (51.5) | 102,290,494 (51.5) | 11,868 (44.6) |
| Medicaid | 48,224,743 (24.3) | 48,216,682 (24.3) | 8,061 (30.3) |
| Medicare | 47,072,067 (23.7) | 47,065,492 (23.7) | 6,575 (24.7) |
| Other/Unknown | 1,020,732 (0.51) | 1,020,631 (0.51) | 101 (0.38) |

**Table 1.** Pooled Demographic and Hospital Characteristics Overall and by Retained Surgical Item Status, 2016 to 2023 (continued)
| Characteristic | Overall,<br>N = 198,619,904 | No RSI,<br>N = 198,593,299 | RSI,<br>N = 26,605 |
| --- | --- | --- | --- |
| Race/Ethnicity, No. (%) |  |  |  |
| Asian Or Pacific Islander | 6,715,261 (3.38) | 6,714,598 (3.38) | 663 (2.49) |
| Black Or African American | 22,616,978 (11.4) | 22,613,245 (11.4) | 3,733 (14.0) |
| Hispanic Or Latino | 26,041,795 (13.1) | 26,038,422 (13.1) | 3,373 (12.7) |
| White | 98,658,630 (49.7) | 98,645,099 (49.7) | 13,531 (50.9) |
| Other | 8,020,108 (4.04) | 8,018,903 (4.04) | 1,205 (4.53) |
| Unknown | 36,567,132 (18.4) | 36,563,032 (18.4) | 4,100 (15.4) |
| Hospital Region, No. (%) |  |  |  |
| Midwest | 41,550,563 (20.9) | 41,544,461 (20.9) | 6,102 (22.9) |
| Northeast | 42,038,948 (21.2) | 42,033,348 (21.2) | 5,600 (21.0) |
| South | 64,145,224 (32.3) | 64,136,988 (32.3) | 8,236 (31.0) |
| West | 33,274,593 (16.8) | 33,270,425 (16.8) | 4,168 (15.7) |
| Unknown | 17,610,576 (8.87) | 17,608,077 (8.87) | 2,499 (9.39) |
| Hospital Type, No. (%) |  |  |  |
| Academic | 56,003,815 (28.2) | 55,992,892 (28.2) | 10,923 (41.1) |
| Community | 121,918,866 (61.4) | 121,905,027 (61.4) | 13,839 (52.0) |
| Unknown | 20,697,223 (10.4) | 20,695,380 (10.4) | 1,843 (6.93) |
| Surgery Year, No. (%) |  |  |  |
| 2016 | 17,901,855 (9.01) | 17,898,931 (9.01) | 2,924 (11.0) |
| 2017 | 22,738,179 (11.4) | 22,734,827 (11.4) | 3,352 (12.6) |
| 2018 | 24,929,930 (12.6) | 24,926,434 (12.6) | 3,496 (13.1) |
| 2019 | 26,419,982 (13.3) | 26,416,505 (13.3) | 3,477 (13.1) |
| 2020 | 24,244,424 (12.2) | 24,241,165 (12.2) | 3,259 (12.2) |
| 2021 | 28,818,894 (14.5) | 28,814,985 (14.5) | 3,909 (14.7) |
| 2022 | 29,119,448 (14.7) | 29,115,897 (14.7) | 3,551 (13.3) |
| 2023 | 24,447,192 (12.3) | 24,444,555 (12.3) | 2,637 (9.91) |
| Elixhauser Score |  |  |  |
| Mean (SD) | 3.7 (8.5) | 3.7 (8.5) | 7.4 (11.4) |
| Median (IQR) | 0 (0, 6) | 0 (0, 6) | 4 (0, 13) |
| Min, Max | -19, 82 | -19, 82 | -18, 70 |

**Table 1.** Pooled Demographic and Hospital Characteristics Overall and by Retained Surgical Item Status, 2016 to 2023 (continued)
| Characteristic | Overall,<br>N = 198,619,904 | No RSI,<br>N = 198,593,299 | RSI,<br>N = 26,605 |
| --- | --- | --- | --- |
| Elixhauser Conditions, No. (%) |  |  |  |
| Congestive Heart Failure | 15,440,295 (7.77) | 15,436,808 (7.77) | 3,487 (13.1) |
| Cardiac Arrhythmia | 30,521,864 (15.4) | 30,514,566 (15.4) | 7,298 (27.4) |
| Valvular Disease | 13,357,255 (6.73) | 13,354,142 (6.72) | 3,113 (11.7) |
| Pulmonary Circulation Disorders | 5,572,599 (2.81) | 5,571,078 (2.81) | 1,521 (5.72) |
| Vascular Disease | 20,893,225 (10.5) | 20,888,544 (10.5) | 4,681 (17.6) |
| Hypertension - Uncomplicated | 61,811,369 (31.1) | 61,802,254 (31.1) | 9,115 (34.3) |
| Hypertension - Complicated | 20,341,578 (10.2) | 20,337,422 (10.2) | 4,156 (15.6) |
| Paralysis | 2,580,278 (1.30) | 2,579,454 (1.30) | 824 (3.10) |
| Other Neurological Disorders | 11,632,099 (5.86) | 11,628,976 (5.86) | 3,123 (11.7) |
| Chronic Pulmonary Disease | 33,232,649 (16.7) | 33,226,419 (16.7) | 6,230 (23.4) |
| Diabetes - Uncomplicated | 11,846,575 (5.96) | 11,844,812 (5.96) | 1,763 (6.63) |
| Diabetes - Complicated | 29,950,323 (15.1) | 29,945,829 (15.1) | 4,494 (16.9) |
| Hypothyroidism | 23,649,570 (11.9) | 23,645,927 (11.9) | 3,643 (13.7) |
| Renal Failure | 18,756,375 (9.44) | 18,752,756 (9.44) | 3,619 (13.6) |
| Liver Disease | 15,565,750 (7.84) | 15,562,326 (7.84) | 3,424 (12.9) |
| Peptic Ulcer Disease | 3,544,893 (1.78) | 3,544,204 (1.78) | 689 (2.59) |
| AIDS/HIV | 927,565 (0.47) | 927,355 (0.47) | 210 (0.79) |
| Lymphoma | 1,775,237 (0.89) | 1,774,891 (0.89) | 346 (1.30) |
| Metastatic Cancer | 5,322,119 (2.68) | 5,320,532 (2.68) | 1,587 (5.97) |
| Solid Tumor Without Metastasis | 14,251,172 (7.18) | 14,248,285 (7.17) | 2,887 (10.9) |
| Rheumatoid Arthritis/Collagen Vascular Diseases | 9,077,089 (4.57) | 9,075,516 (4.57) | 1,573 (5.91) |
| Coagulopathy | 9,663,993 (4.87) | 9,660,956 (4.86) | 3,037 (11.4) |
| Obesity | 41,229,735 (20.8) | 41,222,392 (20.8) | 7,343 (27.6) |
| Weight Loss | 9,495,868 (4.78) | 9,492,921 (4.78) | 2,947 (11.1) |
| Fluid and Electrolyte Disorders | 23,895,427 (12.0) | 23,888,178 (12.0) | 7,249 (27.2) |
| Blood Loss Anemia | 3,879,424 (1.95) | 3,878,419 (1.95) | 1,005 (3.78) |
| Iron Deficiency Anemia | 14,814,361 (7.46) | 14,811,233 (7.46) | 3,128 (11.8) |
| Alcohol Abuse | 6,951,344 (3.50) | 6,949,869 (3.50) | 1,475 (5.54) |
| Drug Abuse | 8,596,913 (4.33) | 8,594,631 (4.33) | 2,282 (8.58) |

**Table 1.** Pooled Demographic and Hospital Characteristics Overall and by Retained Surgical Item Status, 2016 to 2023 (continued)
| Characteristic | Overall,<br>N = 198,619,904 | No RSI,<br>N = 198,593,299 | RSI,<br>N = 26,605 |
| --- | --- | --- | --- |
| Psychoses | 2,389,719 (1.20) | 2,389,125 (1.20) | 594 (2.23) |
| Depression | 34,701,583 (17.5) | 34,695,009 (17.5) | 6,574 (24.7) |

Abbreviations: AIDS = Acquired Immune Deficiency Syndrome, HIV = Human Immunodeficiency Virus, IQR = Inter Quartile Range, RSI = Retained Surgical Item, SD = Standard Deviation

RSI incidence pooled across 2016 to 2023 was 1.34 (95% CI: 1.32 to 1.36) per 10,000 surgical procedures. See **Table 2** for additional details and surgical category-stratified estimates. The following category-stratified incidence estimates were greater than 2.0 per 10,000 surgeries (descending order): exploratory laparotomy, transplant surgery, diagnostic laparoscopy, neurological and spinal surgery, oral and maxillofacial surgery, and vascular surgery. Annual RSI incidence (**Figure 1**) decreased from 1.63 (95% CI: 1.58 to 1.70) in 2016 to 1.08 (95% CI: 1.04 to 1.12) per 10,000 surgeries in 2023, with a visible plateau in this trend in 2020 and 2021 before continuing to decline through 2023.

**Figure 1.**
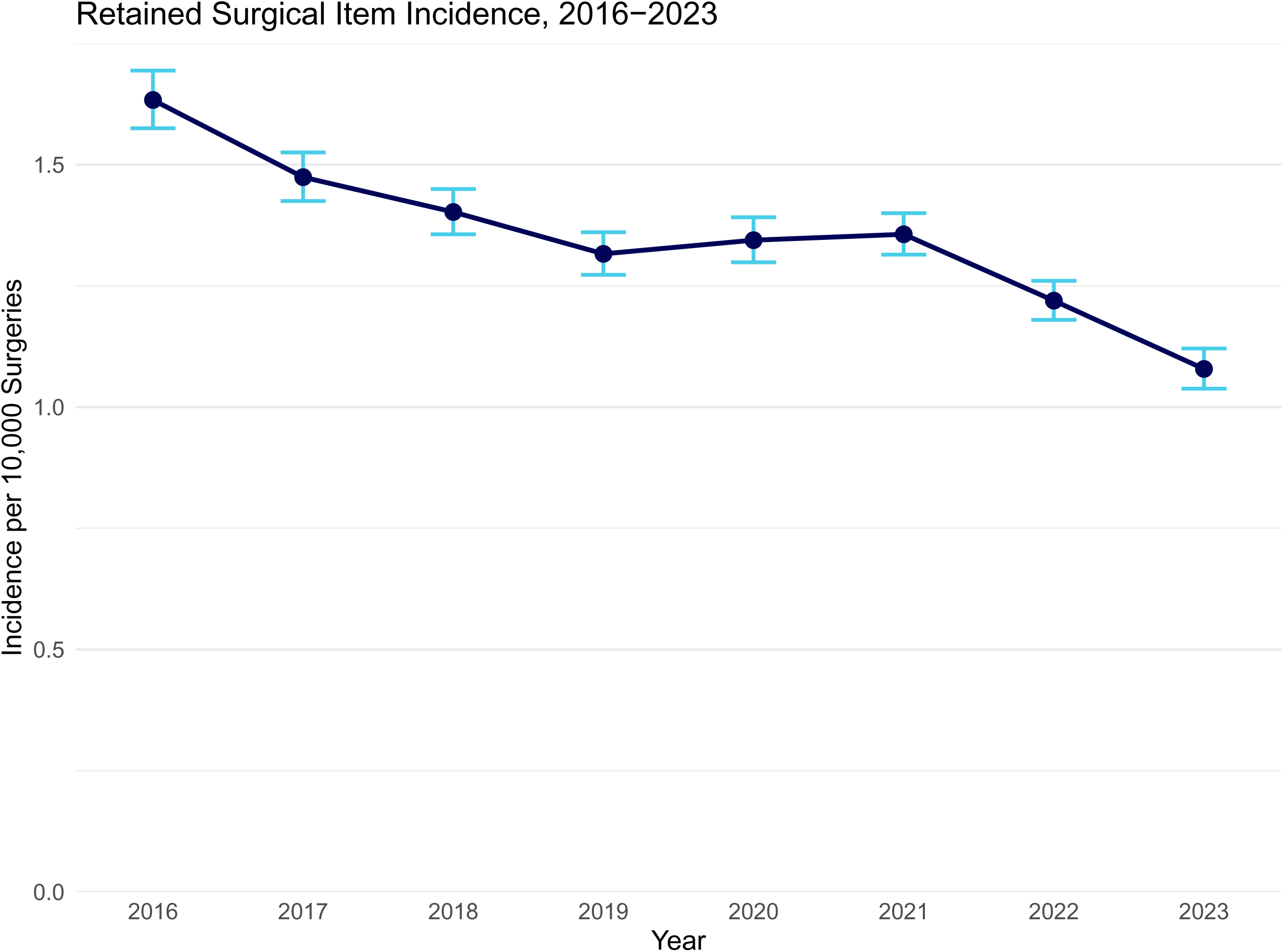
Annual Retained Surgical Item Incidence, 2016 to 2023. Connected dark blue points represent point estimates, light blue bars represent 95% confidence intervals.

**Table 2.** Overall and Surgery Category-Stratified Retained Surgical Item Incidence, 2016 to 2023.

| Characteristic | Number of Incident Events | Number of Surgeries | Retained Surgical Item Incidence per 10,000 Surgeries |  |
| --- | --- | --- | --- | --- |
|  |  |  | Point Estimate | 95% Confidence Interval |
| All Surgeries | 26,605 | 198,619,904 | 1.34 | 1.32 to 1.36 |
| Surgery Category<sup>‡</sup> |  |  |  |  |
| Breast Surgery | 524 | 11,839,759 | 0.44 | 0.41 to 0.48 |
| Cardiac Surgery | 857 | 5,202,662 | 1.65 | 1.54 to 1.76 |
| Diagnostic Laparoscopy | 181 | 241,059 | 7.51 | 6.47 to 8.71 |
| Endocrine Surgery | 46 | 503,282 | 0.91 | 0.68 to 1.23 |
| Exploratory Laparotomy | 171 | 51,939 | 32.92 | 28.26 to 38.33 |
| Gynecologic Surgery | 886 | 11,634,963 | 0.76 | 0.71 to 0.81 |
| Hepatopancreatobiliary and Spleen Surgery | 381 | 4,608,945 | 0.83 | 0.75 to 0.92 |
| Hernia Repair | 125 | 1,246,633 | 1.00 | 0.84 to 1.20 |
| Joint Replacement Surgery | 291 | 3,141,282 | 0.93 | 0.82 to 1.04 |
| Neurological and Spinal Surgery | 908 | 3,630,081 | 2.50 | 2.34 to 2.67 |
| Non-Replacement Orthopedic Surgery | 1,431 | 12,611,828 | 1.13 | 1.08 to 1.20 |
| Obstetric Surgery | 1,583 | 12,246,002 | 1.29 | 1.23 to 1.36 |
| Ophthalmologic Surgery | 380 | 14,177,620 | 0.27 | 0.24 to 0.30 |
| Oral and Maxillofacial Surgery | 30 | 135,110 | 2.22 | 1.52 to 3.21 |
| Otolaryngologic Surgery | 196 | 6,823,858 | 0.29 | 0.25 to 0.33 |
| Soft Tissue and Cutaneous Surgery | 4,474 | 41,171,074 | 1.09 | 1.06 to 1.12 |
| Stomach and Intestinal Surgery | 2,895 | 34,771,989 | 0.83 | 0.80 to 0.86 |
| Thoracic Surgery | 441 | 2,521,355 | 1.75 | 1.59 to 1.92 |
| Transplant Surgery | 103 | 88,118 | 11.69 | 9.59 to 14.23 |
| Urologic Surgery | 720 | 7,214,178 | 1.00 | 0.93 to 1.07 |
| Vascular Surgery | 1,454 | 7,171,273 | 2.03 | 1.93 to 2.14 |

<sup>‡</sup> Surgeries occurring on the same day for a patient as a surgery in another category (and any associated incident events) are excluded from category-specific incidence estimates, as determining which surgery category was responsible for the incident event was not possible.

**Figure 2** depicts the state-stratified RSI incidence; for state-stratified values see **eTable 4**. Geographically, the five highest incidences per 10,000 surgeries (descending order) were in DC, Maine, Nebraska, Ohio, and West Virginia; the five lowest (ascending order) were Utah, North Dakota, South Carolina, Vermont, and Wisconsin.

**Figure 2.**
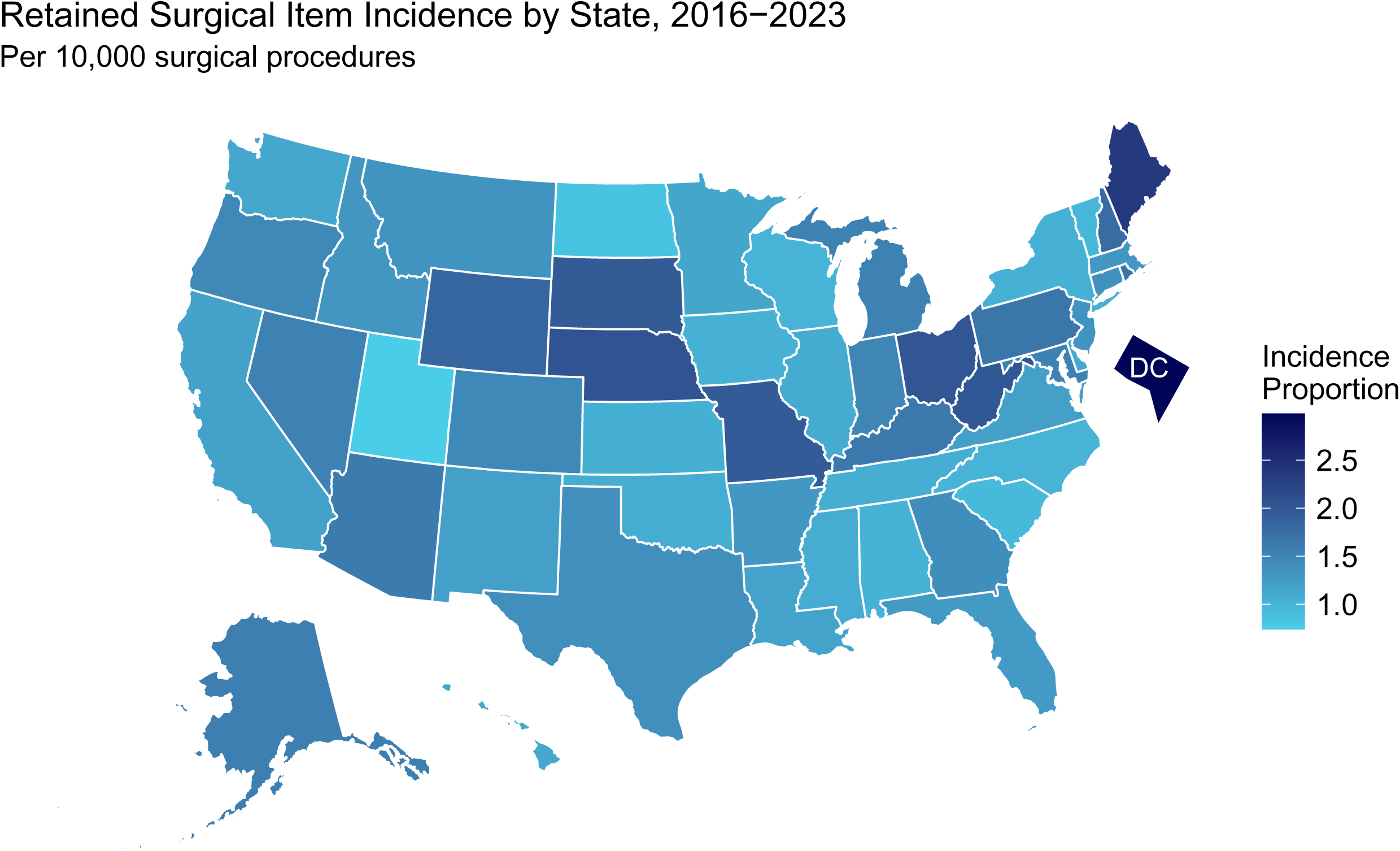
Retained Surgical Item Incidence by State, 2016 to 2023.

The mean time to RSI detection was 30.3 days while median time was 0 days (see **Table 3**), indicating at least half of all RSI events were detected on the same day as surgery. Time to RSI detection estimates stratified by surgery category are presented in **eTable 5**. The distribution of time to RSI detection was similarly skewed for all surgery categories.

**Table 3.**
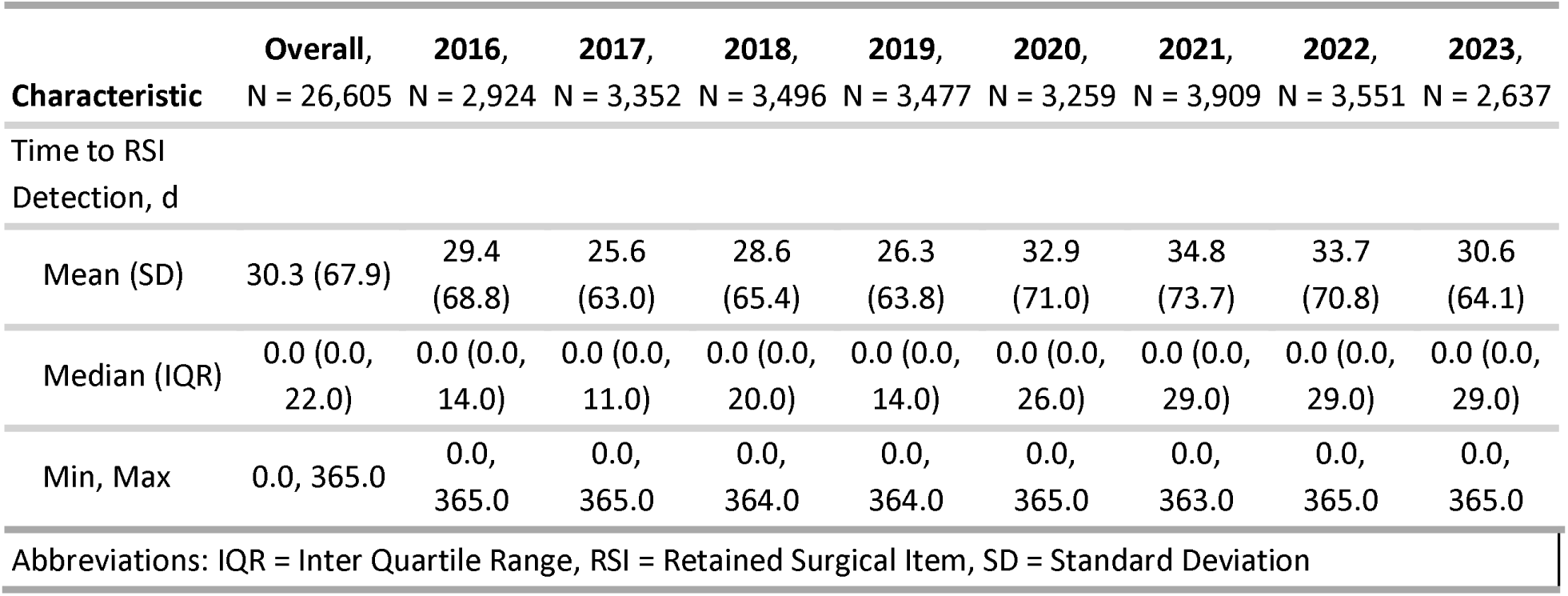
Pooled and Annual Time to Retained Surgical Item Detection, 2016 to 2023.

## Discussion

Our study presents one of the most comprehensive national descriptions of RSIs in the US, leveraging claims data from hundreds of millions of surgeries to quantify incidence trends, demographic patterns, and hospital-level variation. Our findings confirm a persistent but declining RSI incidence from 2016 to 2023. Our pooled incidence estimate is similar to those from other studies, including one study that estimated incidence at 1.3 per 10,000 surgeries using data from 2007 to 2011.^2,3,9^ When examining trends over time, we observed a notable plateau in the decreasing RSI incidence trend that occurred in 2020 and 2021. This coincides with the coronavirus disease 2019 (COVID-19) pandemic, which dramatically altered surgical volumes, procedural priorities, and hospital workflows.^31,32^ During this time, elective operations were deferred, case mix shifted toward emergent surgeries, and hospitals faced system-level strain.^31,32^ The disruption of routine safety checks and the shift to more complex cases with smaller teams may be related to this plateau. Annual RSI incidence continued declining after 2021, suggesting a return to baseline safety practices and operative routines. The relationship between the observed plateau in trend and the COVID-19 pandemic requires formal investigation to substantiate this relationship that is beyond the scope of our study.

The distribution of age, race/ethnicity, sex, and insurance type observed among all surgical cases in our results was consistent with national surgical utilization patterns.^2,12,33–35^ Demographic characteristics among RSI cases were proportional to the distribution among all surgical cases except insurance type, where RSI cases had a higher proportion of Medicaid-insured patients and lower proportion of those commercially-insured. This elevated proportion of Medicaid-insured patients may reflect several interrelated factors. Medicaid-insured patients often experience greater care fragmentation, lower continuity with providers, delayed access to surgical interventions, language barriers, and lower health literacy, which can increase the likelihood of undergoing urgent or emergent procedures, settings that increase the risk of RSIs due to time pressure and complex workflows.^36–38^ Additionally, higher comorbidity burdens, limited access to preoperative optimization, and systemic inequities in care coordination may also contribute to increased medical errors like RSIs in this population.^39–41^

RSI cases also disproportionately occurred at academic hospitals versus community hospitals. This may be due to their role in managing complex surgeries and involving trainees.^3,13,42–45^ Additionally, more rigorous documentation may lead to greater capture of RSIs in claims data compared to community hospitals.^43,46^ Our descriptive study did not account for systematic differences, which require further investigation beyond its scope.

RSIs occurred across a wide range of surgical categories, underscoring there is risk in every surgical field and certain high-risk procedures (e.g., abdominal surgeries) carry higher RSI incidence. This likely relates to the challenge of identifying small or fragmented instruments in large body cavities.^4,12,23^ Exploratory laparotomies had the highest incidence by far, followed by transplant surgery, and diagnostic laparoscopies. These procedures carry inherent complexity and urgency, often involving large surgical teams, extended operative times, and rapid intraoperative decision-making.^9,13^ In exploratory laparotomies, the unpredictable nature of intra-abdominal findings and frequent use of sponges and retractors in deep or obscured spaces heighten the risk of RSI.^4,7,43,47^ Similarly, transplant surgeries involve extensive dissection and reconstruction, which may also increase RSI risk.^48,49^ For these two surgery types in particular, multiple hand-offs and the use of temporary packing materials in these operations may further elevate RSI risk.^22,47^ The high RSI incidence among diagnostic laparoscopies may relate to the confined operative field and challenges in tracking small instruments or materials introduced through ports during rapid assessments.^50^

The number of exploratory laparotomies in our data is likely underestimated due to coding practices, as these procedures often evolve into more definitive procedures and are coded accordingly.^51,52^ Because of the high incidence observed in this category, we expect this misclassification to be differential with respect to RSI risk, leading to overestimation of exploratory laparotomy RSI incidence by retaining RSI cases in the numerator while driving the denominator lower. Procedures in the denominator of other surgery category-stratified estimates may also suffer from misclassification, but do not suffer from the same level of ambiguity as exploratory laparotomy. We estimated category-specific incidence conservatively by excluding surgeries that occurred on the same day in multiple categories to avoid double-counting RSIs.

Patients who experienced an RSI had more comorbidities than those who did not, as reflected in the Elixhauser Comorbidity Index score. Notably, conditions complicating intraoperative management (e.g., coagulopathy, fluid and electrolyte disorders, and obesity) were more common among RSI cases. This suggests that higher physiologic complexity and intraoperative challenges may contribute to increased risk for RSIs.

Time to RSI detection varied notably across different surgical types. Transplant, obstetric, and neurological/spinal surgeries generally demonstrated the most rapid detection, suggesting robust surveillance protocols and heightened clinical awareness in these fields. In contrast, ophthalmologic, otolaryngologic, and gastrointestinal surgeries exhibited longer detection times, which may highlight challenges in symptom recognition or lower immediate clinical suspicion. In support of this, the relatively low incidence of RSIs in ophthalmologic and otolaryngologic surgeries may result in lower immediate clinical suspicion among surgeons, possibly due to the relative rarity of these events. Across surgery categories, most RSIs were detected promptly while a subset of cases were detected much later.

### Limitations

Our study has important limitations. Claims data lack clinical detail, and diagnostic or procedural coding errors may bias estimates. Although the Komodo Research Database provides robust longitudinal coverage, it includes only patients with commercial or government insurance, limiting generalizability to individuals uninsured or treated outside contributing systems. Additionally, assigning an incident RSI to the most recent surgical procedure may result in misclassification of the outcome if a patient has multiple other surgeries where an earlier procedure is the one where the RSI truly occurred. Likewise, by limiting RSI ascertainment based on detection within 365 days post-surgery, further delayed or undetected RSIs were not captured in our study. This could lead to underestimation of RSI incidence, but we expect any such underestimation to be negligible given how rare these cases appear in clinical practice. Missingness of data such as patient demographics, provider characteristics, or hospital-level information may limit the ability to identify disparities in RSI incidence. Hospital classification and geographic assignment relied on available NPI data linked to HCOs or providers. The hierarchical approach we adopted to overcome missing or conflicting data was necessary, but may result in misclassification.

## Conclusions

Pooling surgeries across our study’s 8-year period, RSI incidence was 1.34 per 10,000 surgeries. Despite declining annual incidence from 2016 to 2023, RSIs remain a preventable medical error that continues to occur during surgery. Our study offers a detailed characterization of RSI cases, describing their disproportionate occurrence among Medicaid beneficiaries, patients with greater comorbidity burden, those receiving abdominal and transplant surgeries, and at academic hospitals. The temporary plateau in the decreasing annual RSI incidence trend in 2020 and 2021 highlights that the COVID-19 pandemic may have affected patient safety patterns, which warrants further investigation. Additionally, future research integrating claims data with electronic health records may offer a more complete picture of RSI risk and associated outcomes, enabling targeted quality improvement initiatives. While national organizations have implemented measures to address RSIs, their continued occurrence across diverse surgical contexts highlights the need for sustained, system-wide strategies. Continued vigilance, innovation, and policy support is necessary to further reduce the incidence and downstream burden of RSIs in the US healthcare system.

## Supporting information

eFigure 1, eFigure 2, eMethods

eTable1, eTable 2, eTable 3, eTable 4, eTable 5

## Data Availability

The data used for this study are from the Komodo Research Dataset®, which is built on the Komodo Healthcare Map and is a proprietary data product that requires a data access fee.

## Acknowledgment Section

Author access to data: K. Taylor and P. Kumparatana had full access to all of the data in the study and take responsibility for the integrity of the data and the accuracy of the data analysis.

## Author contributions

*Concept and design:* All authors.

*Acquisition, analysis, or interpretation of data:* All authors.

*Drafting of the manuscript:* Mayan, Kumparatana, Taylor.

*Critical review of the manuscript for important intellectual content:* All authors.

*Statistical analysis:* Kumparatana, Taylor.

*Obtained funding:* N/A.

*Administrative, technical, or material support:* Antony, Taylor.

*Supervision:* Antony, Taylor.

## Potential conflicts of interest

All authors are employees of and receive stock options as part of their compensation from Komodo Health, Inc. No other disclosures were reported.

## Funding/Support

This work was supported by Komodo Health, Inc. via the authors employment, data access, and cloud computation time. The views expressed in this study are those of the authors and may not reflect those of Komodo Health, Inc.

## Role of the Funder/Sponsor

Outside of the authors, Komodo Health, Inc. had no role in the design and conduct of the study; collection, management, analysis, and interpretation of the data; and preparation of the manuscript. Outside of the authors, Komodo Health, Inc. did have a role in review and approval of the manuscript; and decision to submit the manuscript for publication.

## Data Sharing Statement

The data used for this study are from the Komodo Research Dataset, which is built on the Komodo Healthcare Map® and is a proprietary data product that requires a data access fee.

## Notes

### Author Declarations

The Komodo Health data used for this study is de-identified using the Expert Determination de-identification standard as specified by HIPAA (45 CFR Section 164.514(b)(1)). Neither the authors nor Komodo Health have a keycode or crosswalk that can be used to re-identify patients. The use of such de-identified data is not considered human subjects research and does not require ethics approval.

## References

1. Fencl JL. Guideline Implementation: Prevention of Retained Surgical Items. AORN J. 2016;104(1):37–48. doi:10.1016/j.aorn.2016.05.005

2. Elsharydah A, Warmack KO, Minhajuddin A, Moffatt-Bruce SD. Retained surgical items after abdominal and pelvic surgery: Incidence, trend and predictors-observational study. Ann Med Surg. 2016;12:60–64. doi:10.1016/j.amsu.2016.11.006

3. Egorova NN, Moskowitz A, Gelijns A, et al. Managing the Prevention of Retained Surgical Instruments: What Is the Value of Counting? Ann Surg. 2008;247(1):13–18. doi:10.1097/SLA.0b013e3180f633be

4. Hariharan D, Lobo D. Retained surgical sponges, needles and instruments. Ann R Coll Surg Engl. 2013;95(2):87–92. doi:10.1308/003588413X13511609957218

5. Steelman VM, Alasagheirin MH. Assessment of Radiofrequency Device Sensitivity for the Detection of Retained Surgical Sponges in Patients With Morbid Obesity. Arch Surg. 2012;147(10):955. doi:10.1001/archsurg.2012.1556

6. Weprin SA, Meyer D, Li R, et al. Incidence and OR team awareness of “near-miss” and retained surgical sharps: a national survey on United States operating rooms. Patient Saf Surg. 2021;15(1):14. doi:10.1186/s13037-021-00287-5

7. Wallace SC. Retained Surgical Items: Events and Guidelines Revisited. Pa Patient Saf Advis. 2017;14(1):27–35.

8. Snape AJ, Duff J, Gumuskaya O, Inder K, Hutton A. Strategies to prevent inadvertent retained surgical items: An integrative review. J Perioper Nurs. 2022;35(4). doi:10.26550/2209-1092.1196

9. Hempel S, Maggard-Gibbons M, Nguyen DK, et al. Wrong-Site Surgery, Retained Surgical Items, and Surgical Fires: A Systematic Review of Surgical Never Events. JAMA Surg. 2015;150(8):796. doi:10.1001/jamasurg.2015.0301

10. DeWane MP, Kaafarani HMA. Retained Surgical Items: How Do We Get to Zero? Jt Comm J Qual Patient Saf. 2023;49(1):1–2. doi:10.1016/j.jcjq.2022.11.005

11. Joint Commission. Preventing unintended retained foreign objects. Sentin Event Alert. 2013;(51):1–5.

12. Gonzalez-Ojeda A, Rodriguez-Alcantar DA, Arenas-Marquez H, et al. Retained foreign bodies following intra-abdominal surgery. Hepatogastroenterology. 1999;46(26):808–812.

13. Weprin S, Crocerossa F, Meyer D, et al. Risk factors and preventive strategies for unintentionally retained surgical sharps: a systematic review. Patient Saf Surg. 2021;15(1):24. doi:10.1186/s13037-021-00297-3

14. Imputing Allowed Amounts: Development and Validation of an Encounter-Level Allowed Amount Imputation Model. Komodo Health, Inc.; 2023:4. Accessed April 4, 2025. https://knowledge.komodohealth.com/hubfs/Komodo_Imputing_Allowed_Amounts_White_Paper.pdf

15. Health Insurance Portability and Accountability Act of 1996, 45 CFR § 164.514(b)(1) (2025). https://www.ecfr.gov/current/title-45/part-164/subpart-E#p-164.514(b)(1)

16. Healthcare Cost and Utilization Project. Surgery Flag Software for Services and Procedures [computer software]. Version 2024.1. Rockville, MD: Agency for Healthcare Research and Quality; 2024. www.hcup-us.ahrq.gov/toolssoftware/surgeryflags_svcproc/surgeryflagssvc_proc.jsp

17. Patient Safety Indicators Appendix E: Surgical Discharge MS-DRGs, Version V2024. Agency for Healthcare Research and Quality; 2024:1–12. Accessed March 19, 2025. https://qualityindicators.ahrq.gov/measures/PSI_TechSpec

18. Healthcare Cost and Utilization Project. Clinical Classifications Software for Services and Procedures [computer software]. Version 2024.1. Rockville, MD: Agency for Healthcare Research and Quality; 2024. www.hcup-us.ahrq.gov/toolssoftware/ccs_svcsproc/ccssvcproc.jsp

19. Quan H, Sundararajan V, Halfon P, et al. Coding Algorithms for Defining Comorbidities in ICD-9-CM and ICD-10 Administrative Data. Med Care. 2005;43(11):1130–1139. doi:10.1097/01.mlr.0000182534.19832.83

20. Van Walraven C, Austin PC, Jennings A, Quan H, Forster AJ. A Modification of the Elixhauser Comorbidity Measures Into a Point System for Hospital Death Using Administrative Data. Med Care. 2009;47(6):626–633. doi:10.1097/MLR.0b013e31819432e5

21. Patient Safety Indicator 05 (PSI 05) Retained Surgical Item or Unretrieved Device Fragment Count, Version V2024. Agency for Healthcare Research and Quality; 2024:2–3. Accessed March 19, 2025. https://qualityindicators.ahrq.gov/measures/PSI_TechSpec

22. Hibbert PD, Thomas MJW, Deakin A, Runciman WB, Carson-Stevens A, Braithwaite J. A qualitative content analysis of retained surgical items: learning from root cause analysis investigations. Int J Qual Health Care J Int Soc Qual Health Care. 2020;32(3):184–189. doi:10.1093/intqhc/mzaa005

23. Zejnullahu VA, Bicaj BX, Zejnullahu VA, Hamza AR. Retained Surgical Foreign Bodies after Surgery. Open Access Maced J Med Sci. 2017;5(1):97–100. doi:10.3889/oamjms.2017.005

24. Wilson EB. Probable Inference, the Law of Succession, and Statistical Inference. J Am Stat Assoc. 1927;22(158):209–212. doi:10.1080/01621459.1927.10502953

25. dbt Core [computer software]. Version 1.5. Philadelphia, PA: dbt Labs, Inc.; 2023. Accessed April 2, 2025. https://docs.getdbt.com/docs/introduction

26. R Core Team. R: A Language and Environment for Statistical Computing [computer software]. Vienna, Austria: R Foundation for Statistical Computing; 2024. https://www.R-project.org/

27. Di Lorenzo P. usmap: US Maps Including Alaska and Hawaii [R package]. Version 0.7.1. 2024. https://CRAN.R-project.org/package=usmap

28. Satorra P, Carmezim J, Pallarès N, Tebé C, Taylor KA. flowchart: Tidy Flowchart Generator [R package]. Version 0.8.0. 2025. https://bruigtp.github.io/flowchart/

29. Sjoberg DD, Whiting K, Curry M, Lavery JA, Larmarange J. Reproducible Summary Tables with the gtsummary Package. R J. 2021;13(1):570. doi:10.32614/RJ-2021-053

30. Wickham H, Averick M, Bryan J, et al. Welcome to the Tidyverse. J Open Source Softw. 2019;4(43):1686. doi:10.21105/joss.01686

31. Moletta L, Pierobon ES, Capovilla G, et al. International guidelines and recommendations for surgery during Covid-19 pandemic: A Systematic Review. Int J Surg Lond Engl. 2020;79:180–188. doi:10.1016/j.ijsu.2020.05.061

32. Al-Jabir A, Kerwan A, Nicola M, et al. Impact of the Coronavirus (COVID-19) pandemic on surgical practice - Part 1. Int J Surg. 2020;79:168–179. doi:10.1016/j.ijsu.2020.05.022

33. Elia C, Schoenfeld C, Bayer O, Ewald C, Reinhart K, Sakr Y. The impact of age on outcome after major surgical procedures. J Crit Care. 2013;28(4):413–420. doi:10.1016/j.jcrc.2012.12.010

34. Strope SA, Sarma A, Ye Z, Wei JT, Hollenbeck BK. Disparities in the use of ambulatory surgical centers: a cross sectional study. BMC Health Serv Res. 2009;9(1):121. doi:10.1186/1472-6963-9-121

35. Haider AH, Scott VK, Rehman KA, et al. Racial Disparities in Surgical Care and Outcomes in the United States: A Comprehensive Review of Patient, Provider, and Systemic Factors. J Am Coll Surg. 2013;216(3):482–492e12. doi:10.1016/j.jamcollsurg.2012.11.014

36. Dualeh SHA, Schaefer SL, Kunnath N, Ibrahim AM, Scott JW. Health Insurance Status and Unplanned Surgery for Access-Sensitive Surgical Conditions. JAMA Surg. 2024;159(4):420. doi:10.1001/jamasurg.2023.7530

37. Schmerler J, Haft M, Nelson S, Srikumaran U, Best MJ. Payer Status and Racial Disparities in Time to Surgery for Emergent Orthopaedic Procedures. J Am Acad Orthop Surg. 2024;32(21):e1121–e1129. doi:10.5435/JAAOS-D-23-01136

38. Kern LM, Seirup JK, Rajan M, Jawahar R, Stuard SS. Fragmented ambulatory care and subsequent emergency department visits and hospital admissions among Medicaid beneficiaries. Am J Manag Care. 2019;25(3):107–112.

39. Gammel JJ, Moore JW, Reis RJ, et al. Medicaid status is independently predictive of increased complications, readmission, and mortality following primary total shoulder arthroplasty. J Shoulder Elbow Surg. 2025;34(5):1368–1376. doi:10.1016/j.jse.2024.08.035

40. Luan-Erfe BM, Erfe JM, DeCaria B, Okocha O. Limited English Proficiency and Perioperative Patient-Centered Outcomes: A Systematic Review. Anesth Analg. 2023;136(6):1096–1106. doi:10.1213/ANE.0000000000006159

41. Roy M, Corkum JP, Urbach DR, et al. Health Literacy Among Surgical Patients: A Systematic Review and Meta-analysis. World J Surg. 2019;43(1):96–106. doi:10.1007/s00268-018-4754-z

42. Punnen S, Taheri S, Chen L, Scott T, Karimuddin A. Comparing resident operative volumes for routine general surgery cases at academic, urban community, and rural training sites. Can J Surg. 2024;67(4):E273–E278. doi:10.1503/cjs.005323

43. Stawicki SP, Evans DC, Cipolla J, et al. Retained Surgical Foreign Bodies: A Comprehensive Review of Risks and Preventive Strategies. Scand J Surg. 2009;98(1):8–17. doi:10.1177/145749690909800103

44. Minami CA, Dahlke AR, Barnard C, et al. Association Between Hospital Characteristics and Performance on the New Hospital-Acquired Condition Reduction Program’s Surgical Site Infection Measures. JAMA Surg. 2016;151(8):777. doi:10.1001/jamasurg.2016.0408

45. Becher RD, Sukumar N, DeWane MP, et al. Regionalization of emergency general surgery operations: A simulation study. J Trauma Acute Care Surg. 2020;88(3):366–371. doi:10.1097/TA.0000000000002543

46. Classen DC, Resar R, Griffin F, et al. ‘Global Trigger Tool’ Shows That Adverse Events In Hospitals May Be Ten Times Greater Than Previously Measured. Health Aff (Millwood). 2011;30(4):581–589. doi:10.1377/hlthaff.2011.0190

47. Gawande AA, Studdert DM, Orav EJ, Brennan TA, Zinner MJ. Risk Factors for Retained Instruments and Sponges after Surgery. N Engl J Med. 2003;348(3):229–235. doi:10.1056/NEJMsa021721

48. Gruttadauria M, Dunn C, Lin J, et al. Patients’ Expectations for Longevity of Kidney Transplant. Prog Transplant. 2019;29(1):48–53. doi:10.1177/1526924818817045

49. Rigamonti D, Rigamonti KH, Rigamonti AS. Retained Foreign Object Signals a Dangerous Atmosphere in the Operating Room. Cureus. Published online March 9, 2025. doi:10.7759/cureus.80301

50. Giorgi M, Schettini G, La Banca L, et al. Prevention and Treatment of Intraoperative Complications During Gynecological Laparoscopic Surgery: Practical Tips and Tricks—A Narrative Review. Adv Ther. 2025;42(5):2089–2117. doi:10.1007/s12325-025-03165-z

51. Mehta A, Lunardi N, Efron DT, et al. Characterizing the underlying diagnoses for exploratory laparotomies to improve risk-adjustment models of postoperative mortality. J Trauma Acute Care Surg. 2019;86(4):664–669. doi:10.1097/TA.0000000000002090

52. Gandle C, Scott FI, Waljee A, Vajravelu RK, Sansgiry S, Hou JK. Development and Validation of an Administrative Codes Algorithm to Identify Abdominal Surgery and Bowel Obstruction in Patients With Inflammatory Bowel Disease. Crohns Colitis 360. 2021;3(1):otab010. doi:10.1093/crocol/otab010

