## Supplementary material for "Retained Surgical Item Incidence in the United States from 2016 to 2023: A Descriptive Study": eFigure 1, eFigure 2, eMethods

**Supplemental Online Content**

**eMethods.**

**eReferences.**

**eFigure 1.** Study design diagram.

**eFigure 2.** Study flow diagram.

**eMethods**

**Hospital Characteristics Ascertainment**

We ascertained the healthcare organization (HCO) associated with each surgical procedure using the related HCO National Provider Identifier (NPI) number. If the procedure was missing a HCO NPI number but the associated provider NPI number was available, then we assigned the surgical procedure to the HCO listed as that provider’s primary affiliation. We assigned each HCOs a hospital type of either academic or community. Academic hospitals were those identified as teaching hospitals by the Centers for Medicare & Medicaid Services or were otherwise identifiable by HCO name as being affiliated with an academic institution (e.g., HCO name including “university”). Community hospitals were those that were not assigned an academic hospital type. If the HCO was unidentifiable (missing both HCO and provider NPI), then we assigned hospital type as unknown. We used a similar approach for ascertaining hospital state (based on HCO NPI and using provider NPI information if HCO NPI information was unavailable). In cases where an individual receiving surgery had more than one procedure on a given date with conflicting geographic information, hospital state was assigned as the state closest to the patient’s state of residence. Geographic distance was calculated using state centroids derived from US Census Bureau 2024 geographic data.^1–3^ If the patient’s state of residence was missing to use in such cases, then the hospital state for the individual’s surgery on that date was assigned at random out of the conflicting hospital state data.

**eReferences**

1.	Walker K. *tigris: Load Census TIGER/Line Shapefiles* [R package]. Version 2.2.1. 2025. https://github.com/walkerke/tigris

2.	Pebesma E. Simple Features for R: Standardized Support for Spatial Vector Data. *R J*. 2018;10(1):439. doi:10.32614/RJ-2018-009

3.	Pebesma E, Bivand R. *Spatial Data Science: With Applications in R*. 1st ed. Chapman and Hall/CRC; 2023. doi:10.1201/9780429459016

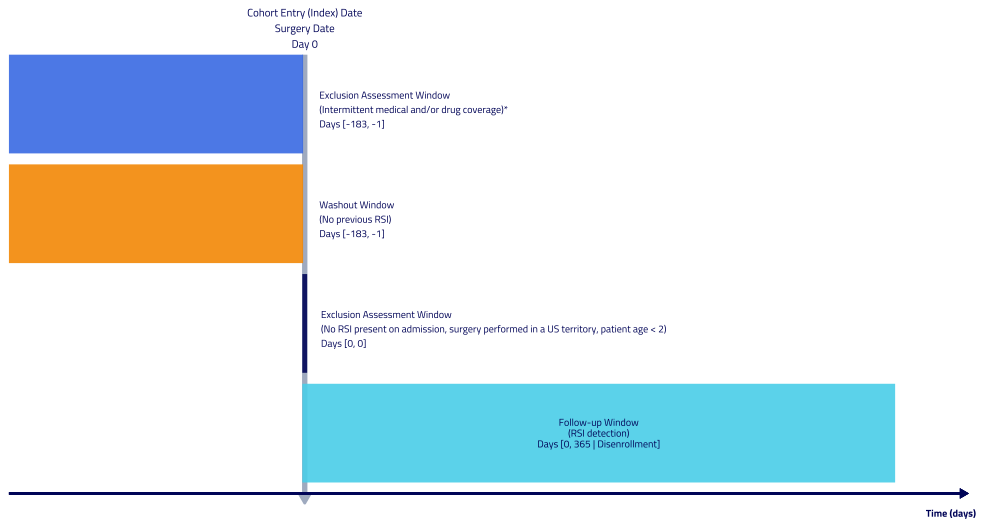

**eFigure 1.** Study design diagram. Data period of June 1, 2015 through December 31, 2024. Eligible surgery events restricted to those in calendar year 2016 through 2023.

*Allowable gaps ≤ 45 days.

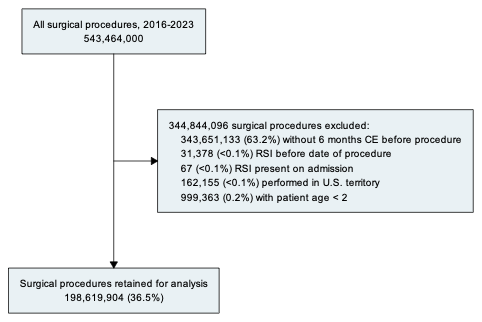

**eFigure 2.** Study flow diagram.
